# A Novel Indicator for COVID-19 Pandemic Assessment and Comparison

**DOI:** 10.1101/2021.10.28.21265596

**Authors:** Kanchan Mukherjee

## Abstract

**Introduction:** In an increasingly globalized world, no country can remain immune to the effect of the COVID-19 pandemic. The pandemic has exposed the need for effective public health surveillance in the interest of global health security. However, current indicators are limited in doing a comparative intercountry assessment and comparison because of variation in testing rates and reporting standards. Hence, this study attempts at addressing this gap.

**Methods:** The study proposes incremental change in cases per testing rate (ICTR) as an indicator for doing cross country comparison of the pandemic progress. The equation for calculating this indicator is explained in this study. This is followed by measuring its strength of association and predictive power for determining change in SARS-CoV-2 cases in five countries (USA, UK, India, Pakistan and Bangladesh).

**Results and discussion:** ICTR was found to have a significantly higher strength of association and predictive power (than the existing indicator-test positivity rate) for determining change in cases over different time periods. Using ICTR, cross country comparison was done for the five countries for15 months to draw deeper insights into the progress of the pandemic.

**Conclusions:** The study finds ICTR to be a suitable indicator for intercountry comparison and intracountry monitoring of the pandemic, which would be useful for global COVID-19 surveillance.

## Background

The ongoing pandemic of coronavirus disease 2019 (COVID-19) caused by the severe acute respiratory syndrome coronavirus 2 (SARS-CoV-2) has brought to focus among many other issues, the importance of public health surveillance. Effective surveillance is dependent on measuring accurate and verifiable indicators so that appropriate preventive and control strategies can be planned and implemented. Surveillance is also required for monitoring long term trends of virus transmission and spread of disease. New cases per day, cumulative total cases, test positivity rates have been used commonly to monitor long term trends in the course of the pandemic. However, these indicators are dependent on testing rates in populations. With a huge variation in testing rates and reporting parameters across different countries, these above indicators are not amenable for comparison because of the confounding effect of testing rates and differential reporting across different countries. In this context, this study explores a new indicator, which is amenable for inter country as well as intra country comparison and monitoring the course of the pandemic.

## Methods

This study is based entirely on secondary data available in the public domain. The world in data COVID-19 database (https://ourworldindata.org/coronavirus) has been used for analysis. Five countries have been used for comparative analysis. These countries are the United States of America (USA), United Kingdom (UK), India, Pakistan and Bangladesh. The study duration was for 15 months starting from June 1, 2020 to August 31, 2021. Three globally reported and recorded existing indicators have been used for analysis. These indicators are:

1. Number of SARS-CoV-2 cases
2. Testing rates (tests/million)
3. Test positivity rate

Since, the number of cases reported are dependent on testing rates and countries have different testing rates, it confounds the accurate comparison of pandemic across different countries. An indicator commonly used for measuring case density and infection spread is test positivity rate (TPR). However, different countries have different reporting standards in terms of testing units like samples tested, persons tested or tests performed which makes intercountry comparison difficult. Hence, the objective of this study was to identify an indicator, which would adjust for confounding factors (testing rates, differential reporting standards) and provide an accurate comparison of intercountry and intracountry pandemic progress with time.

For this purpose, the study proposed that a longitudinal incremental analysis of cases and testing rates would be suitable for measuring the pandemic progress and comparison. Hence, an alternate novel indicator-incremental change in cases per testing rates (ICTR) in equal blocks of time is proposed for comparative assessment of the pandemic. Currently, TPR is a commonly used indicator to assess the severity of the pandemic within a country, and a higher TPR indicates a higher density of infection and vice versa. Hence, this study uses TPR as a comparator. To test whether ICTR is a better measure than TPR for inter country comparison, the study used regression analysis to test the strength of association and predictive power of these two indicators with the change in number of SARS-CoV-2 cases, and compared the respective R, R^2^, F and p-values. TPR and ICTR were the independent variables and change in SARS-CoV-2 cases was the dependent variable. 15 months’ data from two high income countries (HIC) (USA and UK) and three low-middle income countries (LMIC) (India, Pakistan and Bangladesh) have been used for analysis. The 15-month period was divided into five equal blocks of time (3 months each) for this analysis. The conceptual framework for this study is shown in Figure 1.

**Figure 1.**
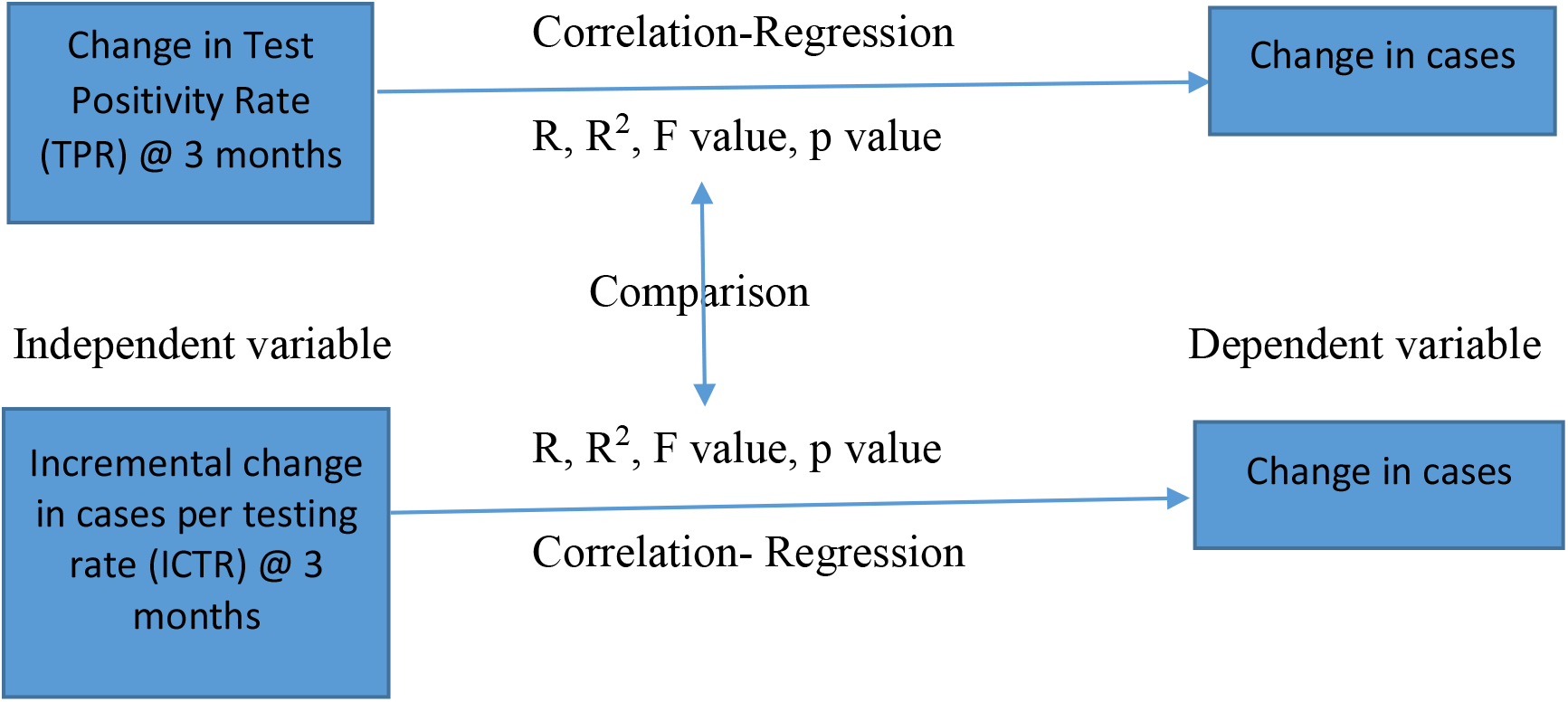
Nomological network of variables analysed in this study (Conceptual framework)

### Calculating the novel indicator – ICTR

ICTR can be calculated using the data on the total SARS-CoV-2 cases and testing rates at any two points in time. The numerator of the indicator is the change in cases during the time period (t) and the denominator is the change in testing rate during the same period (t). Mathematically, if we represent total SARS-CoV-2 cases at beginning of the time period as S1 and the total cases reported at the end of the time period as S2, the numerator will be S2-S1, which is the incremental change in cases during the time period (t). If we represent the testing rate at the beginning of (t) as T1 and testing rate at the end of (t) as T2, the denominator would be T2-T1 (incremental change in testing rate). Hence, the equation for ICTR is as follows:

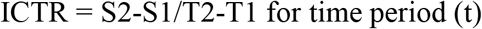

Table 1 shows an application of the ICTR indicator to compare the impact of India’s national lockdown policy on the spread of SARS-CoV-2.

**Table 1.** Impact of national lockdown on SARS-CoV-2 transmission in India (Source: Mukherjee, 2020)

| Time | Day 1 |  | Day 68 |  | Incremental change<br>Cases/testing rate<br>[S2-S1/T2-T1] |
| --- | --- | --- | --- | --- | --- |
|  | Testing rate<br>(Tests/million)<br>(T1) | SARS-CoV-2<br>cases<br>(S1) | Testing rate<br>(Tests/million)<br>(T2) | SARS-CoV-2<br>cases<br>(S2) |  |
| Complete<br>lockdown<br>phase<br>(68 days)<br>(March 25-May 31, 2020) | 18 | 581 | 2708 | 182143 | 67.49 |
| Unlock phase<br>(68 days)<br>(June 1 –<br>August 7.<br>2020) | 2781 | 190535 | 16947 | 2088611 | 133.98 |

As seen from Table 1, India was detecting almost 134 new cases per unit change in testing rate 68 days (t=68 days) after the ‘unlock’; while it was almost half this value (67.49) during the lockdown period (t = 68 days). This indicates that the density of infection was much less during the lockdown period and almost doubled within the first 68 days’ post-lockdown.

## Results and discussion

The results section is structured as follows. In the first section, the study analyses the strength of association (R) and predictive power (R^2^) of the two independent variables (change in TPR and ICTR) to the dependent variable (change in SARS-CoV-2 cases) across equal blocks of three months’ duration in the 15-month study period. These values (R, R^2^, F and p) are then compared with each other as per the conceptual framework shown in Figure 1 to draw inferences. In the second section, the ICTR value is used for empirical comparison among five countries to gain comparative insights into the progress of the pandemic. Tables 2 and 3 show the data on SARS-CoV-2 cases, testing rates, ICTR and TPR for the LMIC and HIC which were used for the regression analysis. The cases, testing rates and TPR data shown in the tables 2 and 3 are from the ‘our world in data’ COVID-19 public database (https://ourworldindata.org/coronavirus)

**Table 2.** Longitudinal comparative analysis of COVID-19 pandemic among three South Asian LMIC countries (15-month period: June 1, 2020 to August 31, 2021)

| **Date / Country** | **1-June, 2020** | **31-Aug** | **1-Sep** | **30-Nov** | **1-Dec** | **28-Feb, 2021** | **1-Mar** | **31-May** | **1-Jun** | **31-Aug** |
| --- | --- | --- | --- | --- | --- | --- | --- | --- | --- | --- |
| **Bangladesh** |  |  |  |  |  |  |  |  |  |  |
| Cases | 49534 | 312996 | 314946 | 464932 | 467225 | 546216 | 546801 | 800540 | 802305 | 150068 |
| Tests/million | 1926 | 9339 | 9413 | 16659 | 16753 | 24300 | 24381 | 35704 | 35813 | 53599 |
| ICTR |  | 35.54 |  | 20.70 |  | 10.47 |  | 22.41 |  | 39.26 |
| TPR | 0.21 | 0.17 | 0.17 | 0.14 | 0.14 | 0.03 | 0.03 | 0.09 | 0.09 | 0.13 |
| **Pakistan** |  |  |  |  |  |  |  |  |  |  |
| Cases | 76398 | 295849 | 296149 | 400482 | 403311 | 581365 | 582528 | 922824 | 924667 | 116368 |
| Tests/million | 2492 | 11639 | 11732 | 24462 | 24644 | 39751 | 39921 | 58710 | 58922 | 78847 |
| ICTR |  | 23.99 |  | 8.20 |  | 11.79 |  | 18.11 |  | 12.00 |
| TPR | 0.24 | 0.02 | 0.02 | 0.07 | 0.07 | 0.03 | 0.03 | 0.04 | 0.04 | 0.07 |
| **India** |  |  |  |  |  |  |  |  |  |  |
| Cases | 198370 | 3691166 | 3769523 | 9462809 | 9499413 | 11112241 | 11124527 | 28175044 | 28307832 | 32810845 |
| Tests/million | 2754 | 30363 | 31093 | 100746 | 101441 | 155181 | 155632 | 247499 | 248880 | 374291 |
| ICTR |  | 126.51 |  | 81.74 |  | 30.01 |  | 185.6 |  | 35.91 |
| TPR | 0.07 | 0.08 | 0.08 | 0.04 | 0.04 | 0.02 | 0.02 | 0.09 | 0.09 | 0.03 |

**Table 3.** Longitudinal comparative analysis of COVID-19 pandemic among two HIC (15-month period: June 1, 2020 to August 31, 2021)

| **Date/**<br>**Country** | **1-June**<br>**2020** | **31-Aug** | **1-Sep** | **30-Nov** | **1-Dec** | **28-Feb**<br>**2021** | **1-Mar** | **31-May** | **1-Jun** | **31-Aug** |
| --- | --- | --- | --- | --- | --- | --- | --- | --- | --- | --- |
| **USA** |  |  |  |  |  |  |  |  |  |  |
| Cases | 1816679 | 6025133 | 6067946 | 13681218 | 13875057 | 28701621 | 28757042 | 33321318 | 33343437 | 39321999 |
| Test/million | 59156 | 274801 | 277527 | 610043 | 615577 | 1039219 | 1042213 | 1353983 | 1356108 | 1605180 |
| ICTR |  | 19.52 |  | 22.90 |  | 35.00 |  | 14.64 |  | 24.00 |
| TPR | 0.043 | 0.048 | 0.048 | 0.102 | 0.105 | 0.052 | 0.052 | 0.024 | 0.025 | 0.099 |
| **UK** |  |  |  |  |  |  |  |  |  |  |
| Cases | 258983 | 338083 | 339415 | 1633736 | 1647233 | 4188827 | 4194289 | 4503231 | 4506333 | 6821356 |
| Test/million | 25392 | 157535 | 159943 | 498690 | 503019 | 1139740 | 1150234 | 2408274 | 2417855 | 3576018 |
| ICTR |  | 0.60 |  | 3.82 |  | 3.99 |  | 0.25 |  | 2.00 |
| TPR | 0.045 | 0.008 | 0.008 | 0.053 | 0.054 | 0.015 | 0.013 | 0.004 | 0.005 | 0.041 |

As seen from Table 2 and 3, there is a variation in testing rates across these five countries and this variation has increased with time. Using the data from the above two tables, regression analysis was conducted based on the conceptual framework shown in Figure 1, which is discussed below.

Table 4 shows the comparative regression analysis results for change in TPR with the change in cases and for ICTR with change in cases during the same time period for the five countries.

**Table 4.** Comparative Regression Analysis

| **▲ TPR vs Change in cases** | **ICTR vs Change in cases** |
| --- | --- |
| **Bangladesh** | **Bangladesh** |
| Multiple R= 0.589 | Multiple R= 0.928 |
| R square = 0.347 | R square = 0.861 |
| Adjusted R square = 0.097 | Adjusted R square = 0.611 |
| F value = 2.13 | F value = 24.81 |
| p value = 0.24 | p value = 0.02* |

| **Pakistan** | **Pakistan** |
| --- | --- |
| Multiple R= 0.593 | Multiple R= 0.949 |
| R square = 0.352 | R square = 0.900 |
| Adjusted R square = 0.102 | Adjusted R square = 0.650 |
| F value = 2.17 | F value = 36.06 |
| p value = 0.24 | p value = 0.009* |
| **India** | **India** |
| Multiple R= 0.88 | Multiple R= 0.93 |
| R square = 0.77 | R square = 0.86 |
| Adjusted R square = 0.52 | Adjusted R square = 0.61 |
| F value = 13.66 | F value = 25.15 |
| p value = 0.05 | p value = 0.015* |
| **USA** | **USA** |
| Multiple R= 0.87 | Multiple R= 0.97 |
| R square = 0.76 | R square = 0.94 |
| Adjusted R square = 0.51 | Adjusted R square = 0.69 |
| F value = 12.41 | F value = 68.58 |
| p value = 0.04 | p value = 0.004* |
| **UK** | **UK** |
| Multiple R= 0.84 | Multiple R= 0.91 |
| R square = 0.71 | R square = 0.83 |
| Adjusted R square = 0.46 | Adjusted R square = 0.58 |
| F value = 9.78 | F value = 19.35 |
| p value = 0.05 | p value = 0.02* |

\*p value significant

Comparing the R, R square, adjusted R square, F and p values from the table above, clearly shows that ICTR has a much stronger association and predictive value (than TPR) for change in SARS-CoV-2 cases across all these countries. Also, ICTR is significantly associated (p < 0.05) with change in cases for all the five countries. Based on the above analysis, the study proposes ICTR as an indicator to provide an accurate comparative assessment of the progress of the pandemic across different countries and within a country. In the next section, the study applies the ICTR for a comparative assessment of these five countries during the 15-month study period.

2. Comparative assessment of pandemic progress using ICTR

Figure 2 shows the comparative assessment of the pandemic for the 15-month study period through the lens of ICTR.

**Figure 2.**
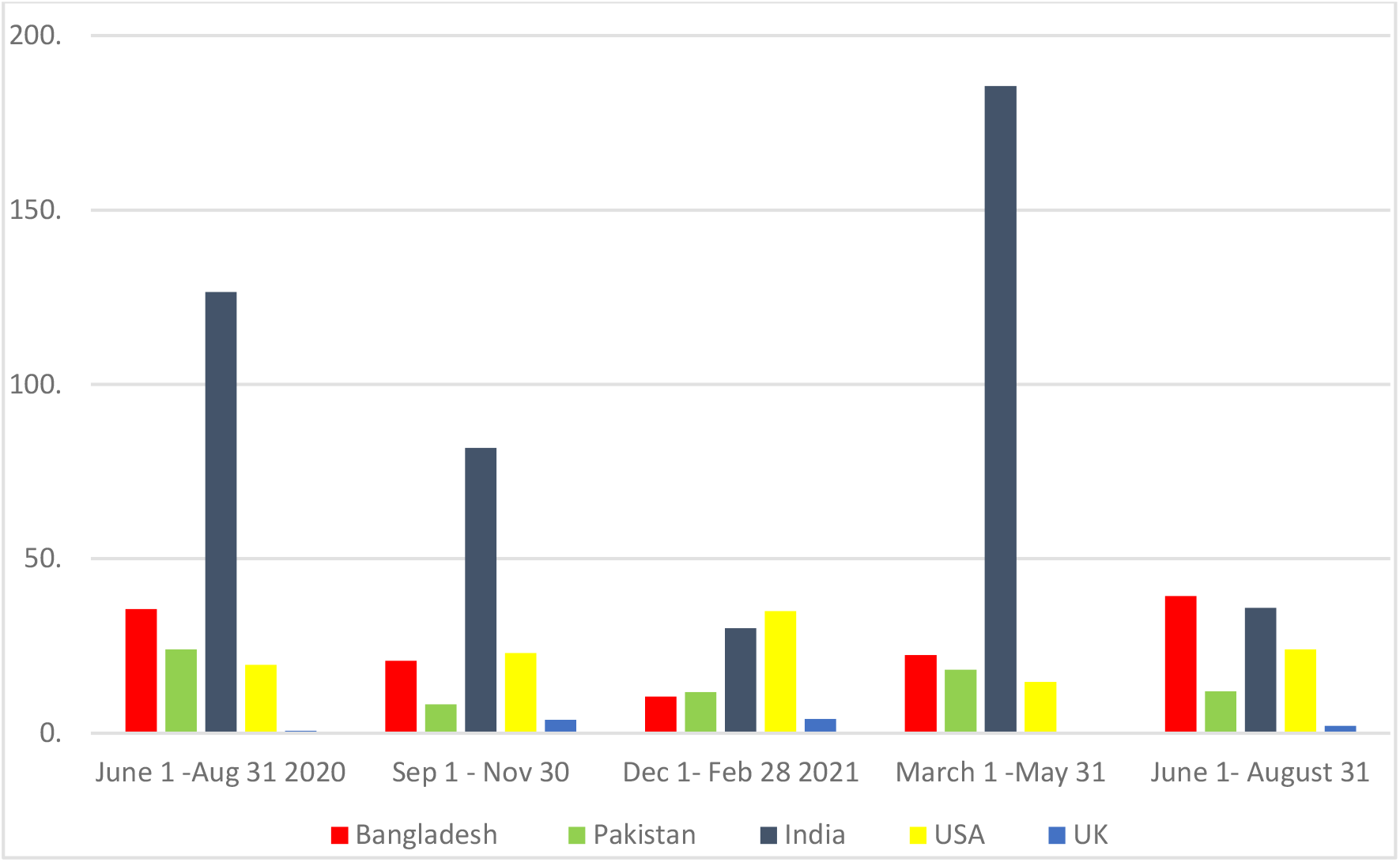
Five country comparison of ICTR (t=3 months)

During the 15-month study period, ANOVA showed highly significant variation in ICTR among these five countries (p = 0.0002). ICTR variance and value has been the maximum for India (4261.5 and 185 respectively), and the least for UK (3.05 and 0.6 respectively). Although, USA has been consistently reporting the highest number of cases during this 15-month period, its ICTR never crossed the 40 mark and UK has had the lowest ICTR consistently among this group indicating a relatively better control of the pandemic among these countries. Among the LMIC, Pakistan has performed relatively well and its ICTR values has been second lowest during three time periods and it has the second lowest variance (38.97) in the group after UK. Bangladesh was able to control the pandemic well till the third period, but after that the ICTR has started to increase and in the last period (June 1 to August 31, 2021) for the first time, Bangladesh had the highest ICTR in this group. This indicates that the pandemic was spreading in Bangladesh during this period, while getting controlled in neighboring India and Pakistan. The ICTR of India crossed the 100 mark twice during this period. The first period (June 1-August 31, 2020) was immediately after the lockdown and the second period (March 1-May 31, 2021) was during the second wave of the pandemic in India. However, after this India’s ICTR has come down below the 40 mark, but still remains higher than the HIC in this group.

Study limitations: The study restricts the analysis to only five countries and hence future studies using ICTR can check its validity and implications for larger set of countries.

## Conclusions

South Asia is one of the most densely populated regions in the world and has comparatively lower testing rates than high income countries like USA and UK, which have much higher testing rates. Since, the detection of the virus is directly associated with testing, comparing just the number of cases across these countries does not provide an accurate comparison as it is confounded by country specific testing rates. Although, TPR gives an indication of the density and spread of infection, given the different reporting standards of testing across countries, TPR is also not a suitable indicator for comparative assessment between countries. In this context, this study proposed an alternate indicator for comparative assessment –ICTR, and statistically tested its strength of association and predictive power in comparison to TPR. It was found that ICTR has a significantly higher association and predictive power for determining change of cases and hence was found suitable for empirical analysis. The empirical analysis of the five countries using ICTR revealed deeper insights into the spread of the pandemic, which was not possible with the conventional indicators.

The advantages of ICTR are as follows:

1. It can be calculated using existing reported data and does not require any additional data collection
2. It is suitable for inter country and intra country comparisons
3. It provides additional insights for understanding the pandemic progress which is useful for planning preventive and control strategies
4. It provides flexibility in deciding the time period (t) for analysis. For example, in this paper, the inter country analysis was done using a t=3 months, while table 1 used ICTR with a t=68 days for analyzing the impact of an intracountry lockdown policy. Hence, future researchers may choose the time period for analysis based on the local context and relevance for their respective study objectives.

Based on the results, this study concludes that ICTR is a feasible indicator to measure and use for comparative assessment of pandemic progress across different countries and within countries, and has implications for pandemic surveillance and control strategies.

## Data Availability

All data produced in the present work are contained in the manuscript

